# Kaiser Permanente National Cross-Vendor Validation of Mammography Artificial Intelligence Computer-Aided Diagnosis Algorithms in a US-Representative Population

**DOI:** 10.64898/2026.09.22.26363619

**Authors:** Vignesh A. Arasu, Tejomay Gadgil, Mark Westley, Albert Pu, Catherine Lee, Cara Smith Gueye, Jason Balkman, Dorota Wisner, Lili Ivansco, Lawrence Kushi

## Abstract

**Background:** Artificial intelligence computer-aided diagnosis (AI CAD) algorithms for screening mammography have shown promise, but independent head-to-head comparisons of commercial algorithms on large, diverse cohorts remain limited.

**Methods:** 786,124 mammography screening exam performed from January 1 2022 to December 31 2023 were identified from electronic medical records. Three FDA-cleared commercial algorithms (A, B, D) and the open-source academic model Mirai (C), were applied to the four standard screening views. Breast cancer within 12 months was ascertained through cancer registry linkage. Algorithms were compared by AUROC; sensitivity, specificity, and PPV at matched AI-positive rates of 5%, 10%, 15%, 20%, 25%; and the proportion of false negatives classified as AI-positive.

**Results:** Of 707,922 examinations with complete data, 4,436 were associated with cancer. Radiologists recalled 6.8% of examinations, with a sensitivity of 67.2%. Algorithm D had the highest AUROC (0.849), followed by algorithm B (0.818), Mirai (0.817), and algorithm A (0.815). Algorithm D also had the highest sensitivity at every AI-positive rate, from 55.3% (95% CI: 54.1, 56.6) at 5% to 77.9% (95% CI: 76.9, 79.0) at 25%, with CIs that did not overlap those of the other algorithms. No algorithm matched radiologist sensitivity at AI-positive rates of 10% or less. Algorithm B flagged a proportion of radiologist false negatives similar to that of algorithm D from 5% through 20% (20.8% vs 20.5% at 5%), despite its lower standalone sensitivity.

**Conclusions:** AI CAD algorithms differed meaningfully in performance on the same large, diverse screening examination cohort, and an open-source academic model performed comparably to two of the three commercial products. Independent local validation at clinically relevant operating points should precede adoption of mammography AI, and algorithm choice and threshold should be matched to the intended workflow.

## Background

Screening mammography reduces breast cancer mortality, but its accuracy in US practice leaves substantial room for improvement. In the Breast Cancer Surveillance Consortium, digital screening mammography had a sensitivity of 86.9%, a specificity of 88.9%, and a recall rate of 11.6%, with wide variation among radiologists.^1^ Most US screening examinations are interpreted by a single radiologist, and first-generation computer-aided detection (CAD), although adopted almost universally after it became reimbursable, did not improve diagnostic accuracy in community practice.^2^

Deep learning–based artificial intelligence (AI) algorithms are a fundamentally different technology. In retrospective studies, AI systems have matched or exceeded the standalone accuracy of radiologists,^3,4^ and prospective and randomized studies in European population-based programs have shown that AI-supported screening can increase cancer detection while substantially reducing radiologist workload.^5–8^ In the randomized MASAI trial, AI-supported screening detected 20% more cancers than standard double reading and reduced screen-reading workload by 44%,^5^ and subsequent analyses reported higher sensitivity and a lower interval cancer rate with AI support.^6^ A nationwide implementation study in Germany similarly found a higher cancer detection rate without an increase in recalls.^8^

Several gaps limit the translation of this evidence to US practice. Nearly all prospective evidence comes from European double-reading programs with biennial screening, low recall rates, and populations that differ from the US screening population in age range, race and ethnicity, and breast density. Systematic reviews have found that most studies of AI accuracy were small, relied on enriched retrospective datasets, and were rarely independent of the algorithm developer, and that few compared multiple algorithms on the same examinations.^9,10^ Where head-to-head comparisons have been performed, performance differed meaningfully among commercial products,^11^ and algorithm performance has been shown to vary with patient characteristics such as age, race and ethnicity, and breast density.^12^ Moreover, vendors report performance at proprietary thresholds, so published sensitivity and specificity values are not directly comparable across products. US health systems considering adoption therefore lack independent, comparative evidence from populations that resemble their own.

Kaiser Permanente (KP) is an integrated health care delivery system that serves approximately 13 million members across eight US regions, with a membership that broadly resembles the populations of the areas it serves^13^ and with screening, pathology, and cancer registry data linked across the care continuum, with breast cancer incidence rates that match national estimates. We previously used KP data to compare mammography AI algorithms for long-term breast cancer risk prediction.^14^ In the present study, we conducted a national, independent, cross-vendor validation of mammography AI CAD algorithms. Our objectives were (a) to compare the standalone performance of three commercial AI CAD algorithms and Mirai, an academic deep learning model,^15^ on the same screening examinations at matched operating points and (b) to evaluate the extent to which each algorithm identified cancers that were missed by radiologists in routine interpretation.

## Methods

### Study Design and Oversight

This retrospective cohort study was approved by the KP institutional review board, which waived the requirement for informed consent. The study is reported in accordance with the Standards for Reporting of Diagnostic Accuracy Studies (STARD) 2015 and the Checklist for Artificial Intelligence in Medical Imaging (CLAIM). Commercial AI developers provided software under research agreements but had no role in study design, data analysis, interpretation, or manuscript preparation, and had no access to cancer outcomes.

### Study Population

We identified screening mammography examinations performed from January 1, 2022, through December 31, 2023. All examinations were two-dimensional full-field digital mammograms and 3D digital breast tomosynthesis were included, and the most recent prior examination. We excluded examinations in that could not be processed by all algorithms. For women with more than one eligible examination, the most recent exam was selected as the index examination.

### Covariates

Demographic and clinical characteristics were obtained from KP electronic health records and mammography reporting systems. These included age at examination; self-reported race and ethnicity; body mass index; age at first live birth; family history of breast cancer in first- and second-degree relatives; history of benign breast biopsy, categorized by the most severe histologic finding (nonproliferative, proliferative without atypia, proliferative with atypia, or lobular carcinoma in situ); Breast Imaging Reporting and Data System (BI-RADS) breast density as assessed by the interpreting radiologist; and availability of a prior mammogram. Race and ethnicity were collected to characterize the study population and its representativeness.

### Outcome Ascertainment

The reference standard was a diagnosis of breast cancer (invasive carcinoma or ductal carcinoma in situ) within 12 months of the index screening examination, ascertained through linkage to KP regional cancer registries, which report to Surveillance, Epidemiology, and End Results (SEER) and state registries, supplemented by pathology records. Examinations without a cancer diagnosis in this interval were classified as cancer-negative; women without cancer had a median follow-up of 3.0 years.

### Radiologist Interpretation

Clinical interpretations were rendered by KP radiologists as part of routine care without AI decision support. Screening examinations assigned a BI-RADS assessment of 0 were considered positive (recalled), and those assigned BI-RADS 1 or 2 were considered negative. Cross-classifying the radiologist assessment with cancer status defined radiologist true-positive, false-negative, true-negative, and false-positive examinations.

### AI Algorithms

We evaluated three commercially available AI CAD algorithms from three developers, designated A, B, and D to preserve vendor anonymity, together with Mirai, a publicly available deep learning model developed to predict future breast cancer risk from screening mammograms.^15^ Mirai was included as an open-source academic comparator; although designed for risk prediction, its 1-year risk output ranks examinations by the likelihood of cancer and can therefore be evaluated with the same measures as detection algorithms. Each algorithm processed the standard screening views of each examination within KP’s secure computing environment and produced a continuous examination-level score. No algorithm was trained or fine-tuned on KP data. The commercial algorithms were all FDA-cleared.

### Operating Points

Because the algorithms produce scores on different scales and vendor-recommended thresholds differ, we compared algorithms at matched operating points. For each algorithm, we identified the score thresholds at which 5%, 10%, 15%, 20%, and 25% of examinations were classified as AI-positive (the AI-positive rate, analogous to a recall rate). Matching on the AI-positive rate holds constant the number of examinations flagged, and therefore the downstream workload and, approximately, the specificity, so that differences among algorithms are expressed as differences in sensitivity. Because thresholds were selected from the score distribution alone, without reference to cancer outcomes, this approach does not optimistically bias the estimated sensitivity. For algorithms with discrete score outputs, we used the threshold that yielded the AI-positive rate closest to, without exceeding, the target.

### Performance Measures

We assessed overall discrimination with the area under the receiver operating characteristic curve (AUROC). At each operating point, we estimated sensitivity (the proportion of cancers classified as AI-positive), specificity (the proportion of cancer-negative examinations classified as AI-negative), and positive predictive value (PPV; the proportion of AI-positive examinations with cancer).

To evaluate complementarity with radiologist interpretation, we calculated the proportion of radiologist false-negative examinations (cancers assessed as BI-RADS 1 or 2) that were AI-positive, which represents cancers the algorithm could potentially rescue if used as an additional reader, and the proportion of radiologist true-negative examinations that were AI-positive, which represents the additional false-positive burden of that use. To evaluate a rule-out scenario in which AI-negative examinations would be removed from the radiologist worklist, we calculated the proportions of AI-negative examinations that were radiologist true-positive examinations (screen-detected cancers that would be forgone) and radiologist false-positive examinations (false-positive recalls that would be avoided).

### Statistical Analysis

We summarized cohort characteristics with counts and percentages, or medians and interquartile ranges (IQRs). We calculated 95% CIs for sensitivity using the normal approximation and for AUROC using the DeLong method. Analyses were performed using R version 4.6.1 (R Foundation for Statistical Computing)].

## Results

### Cohort Characteristics

The cohort included 786,124 screening examinations, of which 6,314 (8.0 per 1,000) were associated with a breast cancer diagnosis (Table 1). Distributions of characteristics are reported for the 707,922 examinations (4,436 cancers) with complete covariate and assessment data. Three-quarters of examinations (75.0%) were in women aged 50 years or older, and women with cancer were older than those without cancer (43.5% vs 29.0% aged ≥65 years).

**Table 1.**
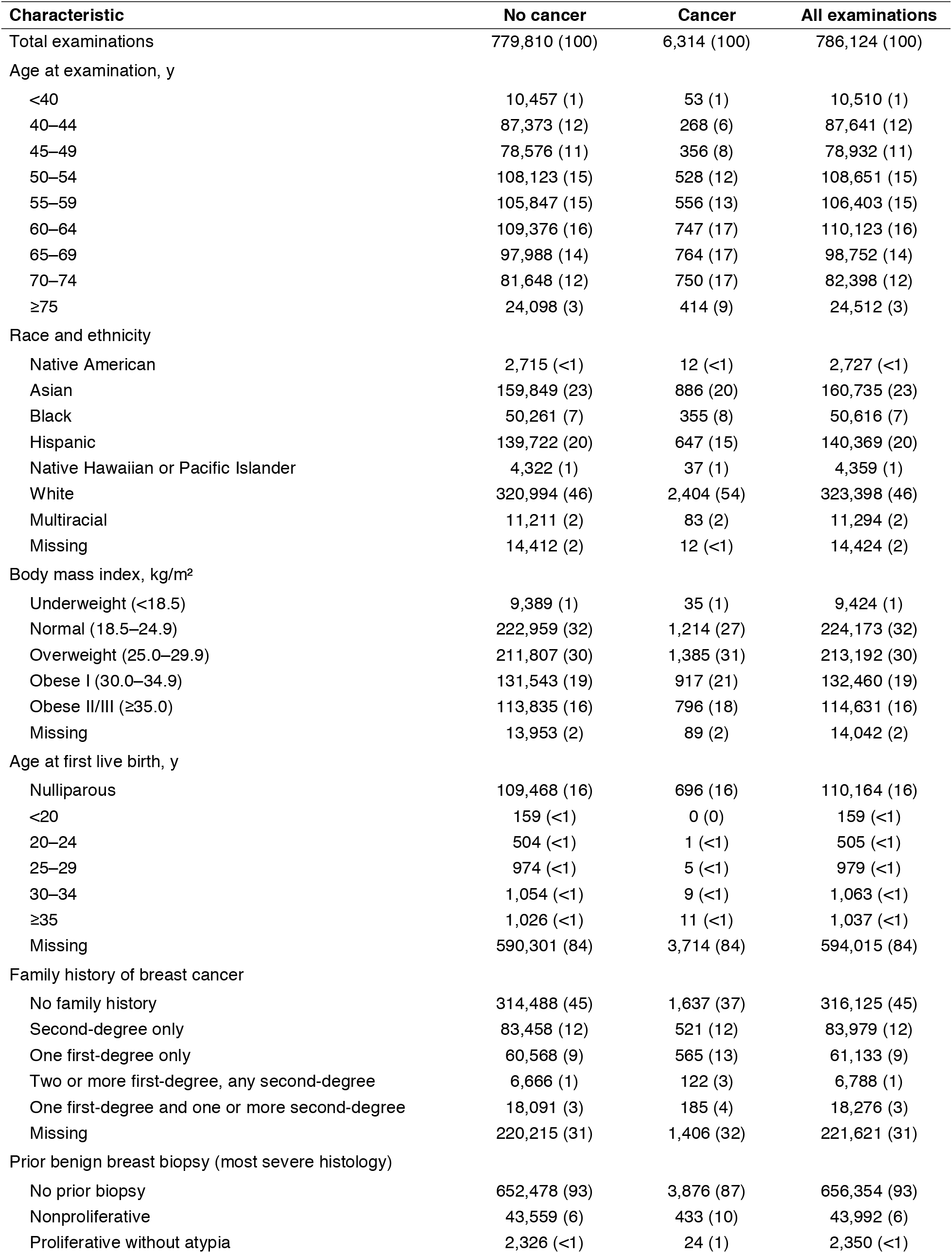
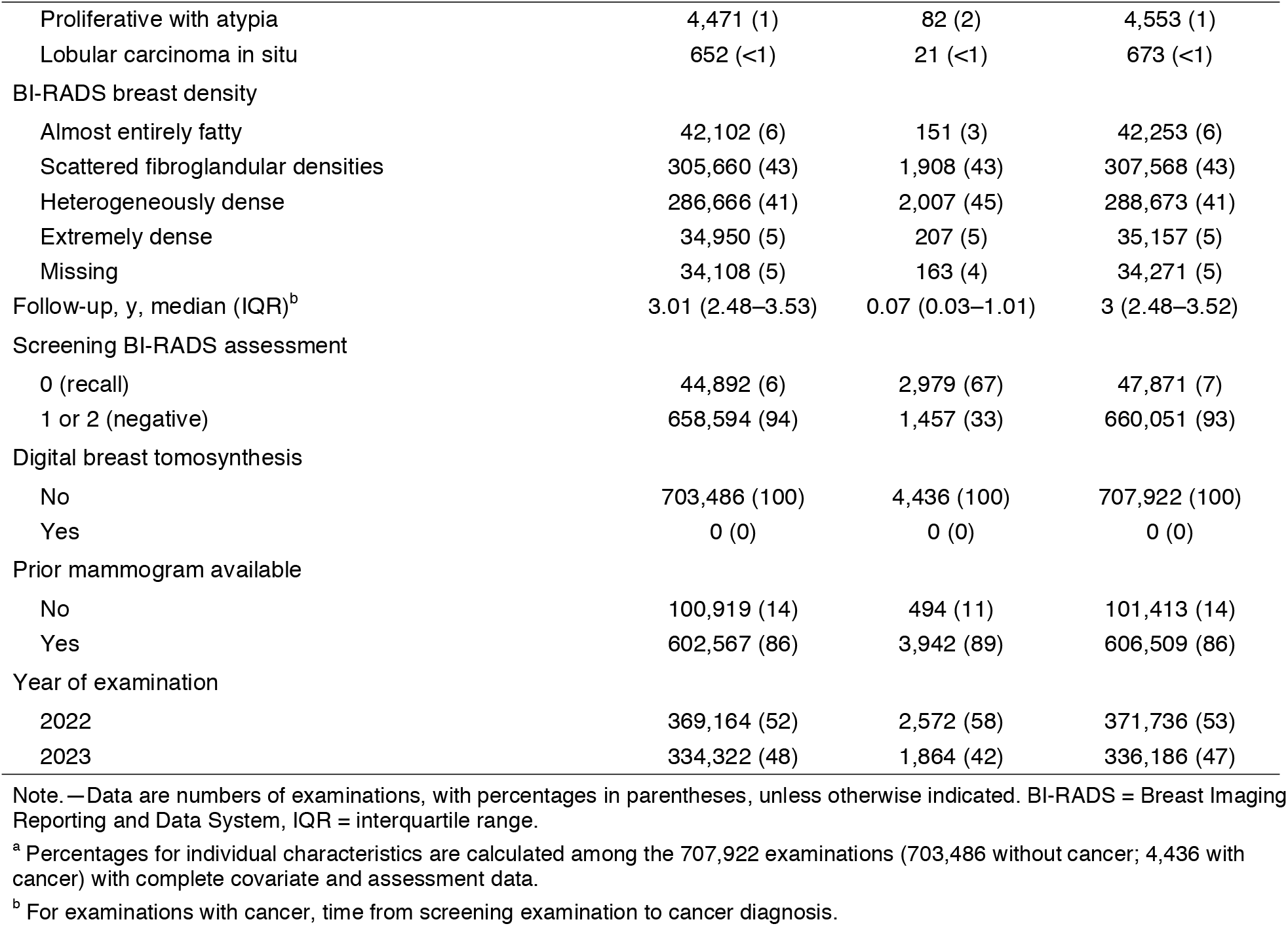
Characteristics of the Study Cohort, by Breast Cancer Status^a^.

| Characteristic | No cancer | Cancer | All examinations |
| --- | --- | --- | --- |
| Total examinations | 779,810 (100) | 6,314 (100) | 786,124 (100) |
| Age at examination, y |  |  |  |
| <40 | 10,457 (1) | 53 (1) | 10,510 (1) |
| 40–44 | 87,373 (12) | 268 (6) | 87,641 (12) |
| 45–49 | 78,576 (11) | 356 (8) | 78,932 (11) |
| 50–54 | 108,123 (15) | 528 (12) | 108,651 (15) |
| 55–59 | 105,847 (15) | 556 (13) | 106,403 (15) |
| 60–64 | 109,376 (16) | 747 (17) | 110,123 (16) |
| 65–69 | 97,988 (14) | 764 (17) | 98,752 (14) |
| 70–74 | 81,648 (12) | 750 (17) | 82,398 (12) |
| ≥75 | 24,098 (3) | 414 (9) | 24,512 (3) |
| Race and ethnicity |  |  |  |
| Native American | 2,715 (<1) | 12 (<1) | 2,727 (<1) |
| Asian | 159,849 (23) | 886 (20) | 160,735 (23) |
| Black | 50,261 (7) | 355 (8) | 50,616 (7) |
| Hispanic | 139,722 (20) | 647 (15) | 140,369 (20) |
| Native Hawaiian or Pacific Islander | 4,322 (1) | 37 (1) | 4,359 (1) |
| White | 320,994 (46) | 2,404 (54) | 323,398 (46) |
| Multiracial | 11,211 (2) | 83 (2) | 11,294 (2) |
| Missing | 14,412 (2) | 12 (<1) | 14,424 (2) |
| Body mass index, kg/m<sup>2</sup> |  |  |  |
| Underweight (<18.5) | 9,389 (1) | 35 (1) | 9,424 (1) |
| Normal (18.5–24.9) | 222,959 (32) | 1,214 (27) | 224,173 (32) |
| Overweight (25.0–29.9) | 211,807 (30) | 1,385 (31) | 213,192 (30) |
| Obese I (30.0–34.9) | 131,543 (19) | 917 (21) | 132,460 (19) |
| Obese II/III (≥35.0) | 113,835 (16) | 796 (18) | 114,631 (16) |
| Missing | 13,953 (2) | 89 (2) | 14,042 (2) |
| Age at first live birth, y |  |  |  |
| Nulliparous | 109,468 (16) | 696 (16) | 110,164 (16) |
| <20 | 159 (<1) | 0 (0) | 159 (<1) |
| 20–24 | 504 (<1) | 1 (<1) | 505 (<1) |
| 25–29 | 974 (<1) | 5 (<1) | 979 (<1) |
| 30–34 | 1,054 (<1) | 9 (<1) | 1,063 (<1) |
| ≥35 | 1,026 (<1) | 11 (<1) | 1,037 (<1) |
| Missing | 590,301 (84) | 3,714 (84) | 594,015 (84) |
| Family history of breast cancer |  |  |  |
| No family history | 314,488 (45) | 1,637 (37) | 316,125 (45) |
| Second-degree only | 83,458 (12) | 521 (12) | 83,979 (12) |
| One first-degree only | 60,568 (9) | 565 (13) | 61,133 (9) |
| Two or more first-degree, any second-degree | 6,666 (1) | 122 (3) | 6,788 (1) |
| One first-degree and one or more second-degree | 18,091 (3) | 185 (4) | 18,276 (3) |
| Missing | 220,215 (31) | 1,406 (32) | 221,621 (31) |
| Prior benign breast biopsy (most severe histology) |  |  |  |
| No prior biopsy | 652,478 (93) | 3,876 (87) | 656,354 (93) |
| Nonproliferative | 43,559 (6) | 433 (10) | 43,992 (6) |
| Proliferative without atypia | 2,326 (<1) | 24 (1) | 2,350 (<1) |

| **Characteristic** | **No cancer** | **Cancer** | **All examinations** |
| --- | --- | --- | --- |
| Proliferative with atypia | 4,471 (1) | 82 (2) | 4,553 (1) |
| Lobular carcinoma in situ | 652 (<1) | 21 (<1) | 673 (<1) |
| BI-RADS breast density |  |  |  |
| Almost entirely fatty | 42,102 (6) | 151 (3) | 42,253 (6) |
| Scattered fibroglandular densities | 305,660 (43) | 1,908 (43) | 307,568 (43) |
| Heterogeneously dense | 286,666 (41) | 2,007 (45) | 288,673 (41) |
| Extremely dense | 34,950 (5) | 207 (5) | 35,157 (5) |
| Missing | 34,108 (5) | 163 (4) | 34,271 (5) |
| Follow-up, y, median (IQR)<sup>b</sup> | 3.01 (2.48–3.53) | 0.07 (0.03–1.01) | 3 (2.48–3.52) |
| Screening BI-RADS assessment |  |  |  |
| 0 (recall) | 44,892 (6) | 2,979 (67) | 47,871 (7) |
| 1 or 2 (negative) | 658,594 (94) | 1,457 (33) | 660,051 (93) |
| Digital breast tomosynthesis |  |  |  |
| No | 703,486 (100) | 4,436 (100) | 707,922 (100) |
| Yes | 0 (0) | 0 (0) | 0 (0) |
| Prior mammogram available |  |  |  |
| No | 100,919 (14) | 494 (11) | 101,413 (14) |
| Yes | 602,567 (86) | 3,942 (89) | 606,509 (86) |
| Year of examination |  |  |  |
| 2022 | 369,164 (52) | 2,572 (58) | 371,736 (53) |
| 2023 | 334,322 (48) | 1,864 (42) | 336,186 (47) |

Note.—Data are numbers of examinations, with percentages in parentheses, unless otherwise indicated. BI-RADS = Breast Imaging Reporting and Data System, IQR = interquartile range.
<sup>a</sup> Percentages for individual characteristics are calculated among the 707,922 examinations (703,486 without cancer; 4,436 with cancer) with complete covariate and assessment data.
<sup>b</sup> For examinations with cancer, time from screening examination to cancer diagnosis.

The cohort was racially and ethnically diverse: 45.7% of examinations were in White women, 22.7% in Asian women, 19.8% in Hispanic women, 7.1% in Black women, 1.6% in multiracial women, 0.6% in Native Hawaiian or Pacific Islander women, and 0.4% in Native American women. Overall, 34.9% of examinations were in women with obesity, and 45.7% were in women with heterogeneously or extremely dense breasts (49.9% among cancers vs 45.7% among non-cancers). Compared with women without cancer, women with cancer more often had a first-degree family history of breast cancer (19.7% vs 12.1%) and a prior benign breast biopsy (12.6% vs 7.3%). A prior mammogram was available for 85.7% of examinations. Among cancers, the median time from screening examination to diagnosis was 0.07 years (IQR, 0.03– 1.01).

### Radiologist Performance

Radiologists recalled 47,871 of 707,922 examinations (6.8%). Of the 4,436 cancers, 2,979 were recalled, corresponding to a sensitivity of 67.2%, a specificity of 93.6% (658,594 of 703,486), a PPV of 6.2%, and a cancer detection rate of 4.2 per 1,000 examinations.

### Discrimination

Algorithm D had the highest AUROC (0.849), followed by algorithm B (0.818), Mirai (0.817), and algorithm A (0.815) (Figure 1). The receiver operating characteristic curves of algorithms A and B and Mirai largely overlapped, whereas the curve for algorithm D lay above the others across the clinically relevant range of low false-positive fractions.

**Figure 1.**
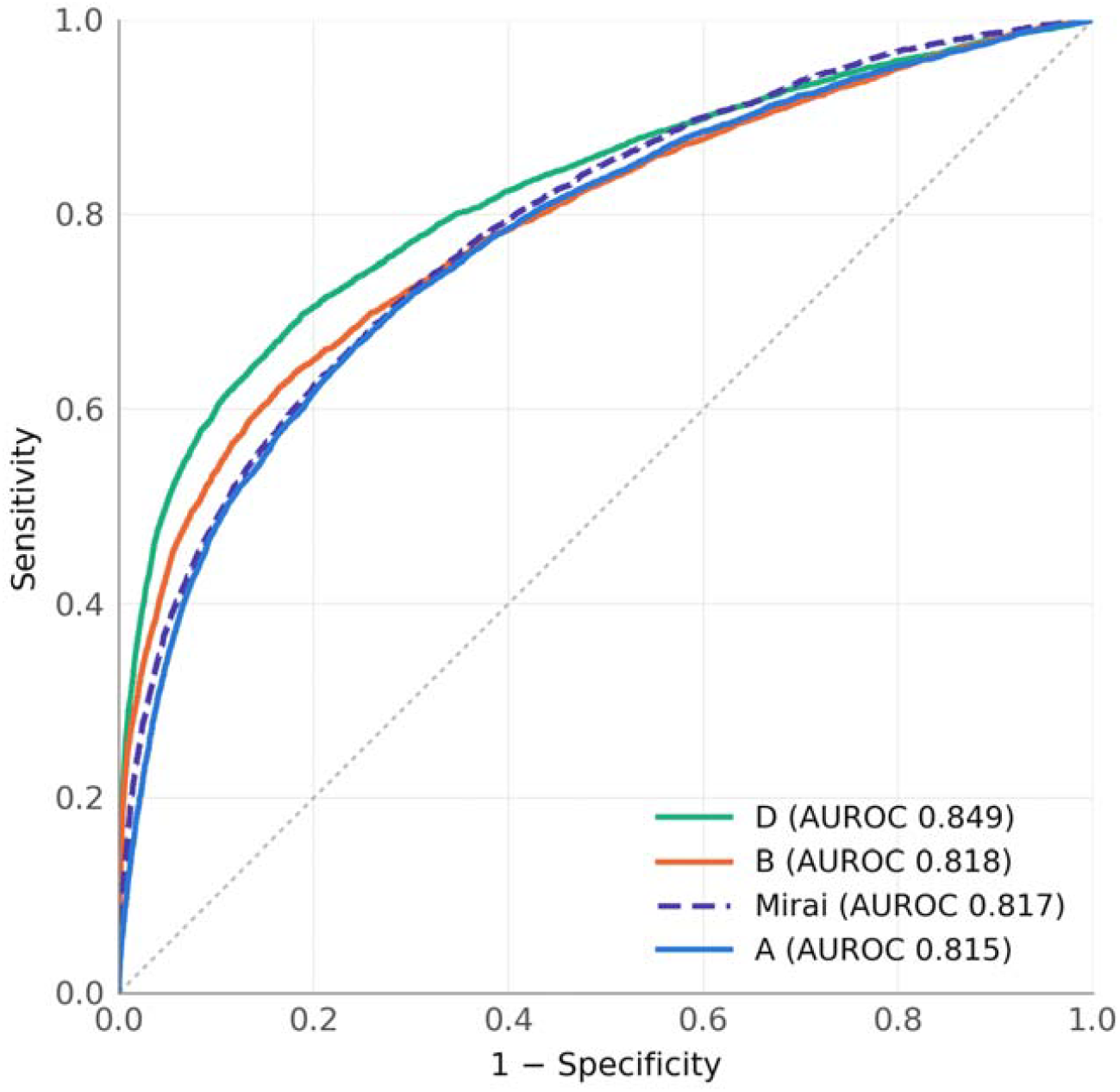
Receiver operating characteristic curves for three commercial AI computer-aided diagnosis algorithms (A, B, and D) and Mirai. The area under the receiver operating characteristic curve (AUROC) for each algorithm is shown in the legend. The dotted diagonal line indicates chance performance.

### Standalone Performance at Matched Operating Points

Sensitivity differed across algorithms at every operating point (Table 2; Figure 2A). At an AI-positive rate of 5%, sensitivity ranged from 42.2% (algorithm A) to 55.3% (algorithm D).

**Table 2.** Standalone Performance of AI Algorithms at Matched AI-Positive Rates.

| Algorithm | Achieved AI-positive rate, % | Sensitivity, % (95% CI) | Specificity, % | PPV, % |
| --- | --- | --- | --- | --- |
| **5% AI-positive rate** |  |  |  |  |
| A | 5.0 | 42.2 (41.0–43.5) | 95.3 | 6.0 |
| B | 5.0 | 47.6 (46.3–49.0) | 95.3 | 6.8 |
| D | 5.0 | 55.3 (54.1–56.6) | 95.4 | 7.9 |
| Mirai | 5.0 | 43.0 (41.7–44.3) | 95.3 | 6.1 |
| **10% AI-positive rate** |  |  |  |  |
| A | 10.0 | 54.0 (52.7–55.3) | 90.3 | 3.8 |
| B | 10.0 | 58.3 (56.9–59.6) | 90.3 | 4.1 |
| D | 10.0 | 64.6 (63.4–65.8) | 90.4 | 4.6 |
| Mirai | 10.0 | 53.9 (52.6–55.3) | 90.3 | 3.8 |
| **15% AI-positive rate** |  |  |  |  |
| A | 15.0 | 62.5 (61.2–63.7) | 85.3 | 3.0 |
| B | 15.0 | 64.4 (63.1–65.6) | 85.4 | 3.1 |
| D | 15.0 | 70.5 (69.4–71.7) | 85.4 | 3.3 |
| Mirai | 15.0 | 61.0 (59.7–62.3) | 85.3 | 2.9 |
| **20% AI-positive rate** |  |  |  |  |
| A | 20.0 | 68.4 (67.2–69.6) | 80.3 | 2.4 |
| B | 20.0 | 68.6 (67.4–69.9) | 80.3 | 2.4 |
| D | 19.9 | 74.2 (73.1–75.3) | 80.5 | 2.6 |
| Mirai | 20.0 | 66.8 (65.6–68.1) | 80.3 | 2.4 |
| **25% AI-positive rate** |  |  |  |  |
| A | 25.0 | 73.1 (72.0–74.3) | 75.3 | 2.1 |
| B | 25.0 | 72.4 (71.2–73.6) | 75.3 | 2.1 |
| D | 25.0 | 77.9 (76.9–79.0) | 75.4 | 2.2 |
| Mirai | 25.0 | 72.1 (71.0–73.3) | 75.3 | 2.1 |

Note.—Thresholds for each algorithm were set to classify the stated percentage of screening examinations as AI-positive. Algorithms A, B, and D are commercial AI computer-aided diagnosis products; Mirai is an open-source academic model. PPV = positive predictive value.

**Figure 2.**
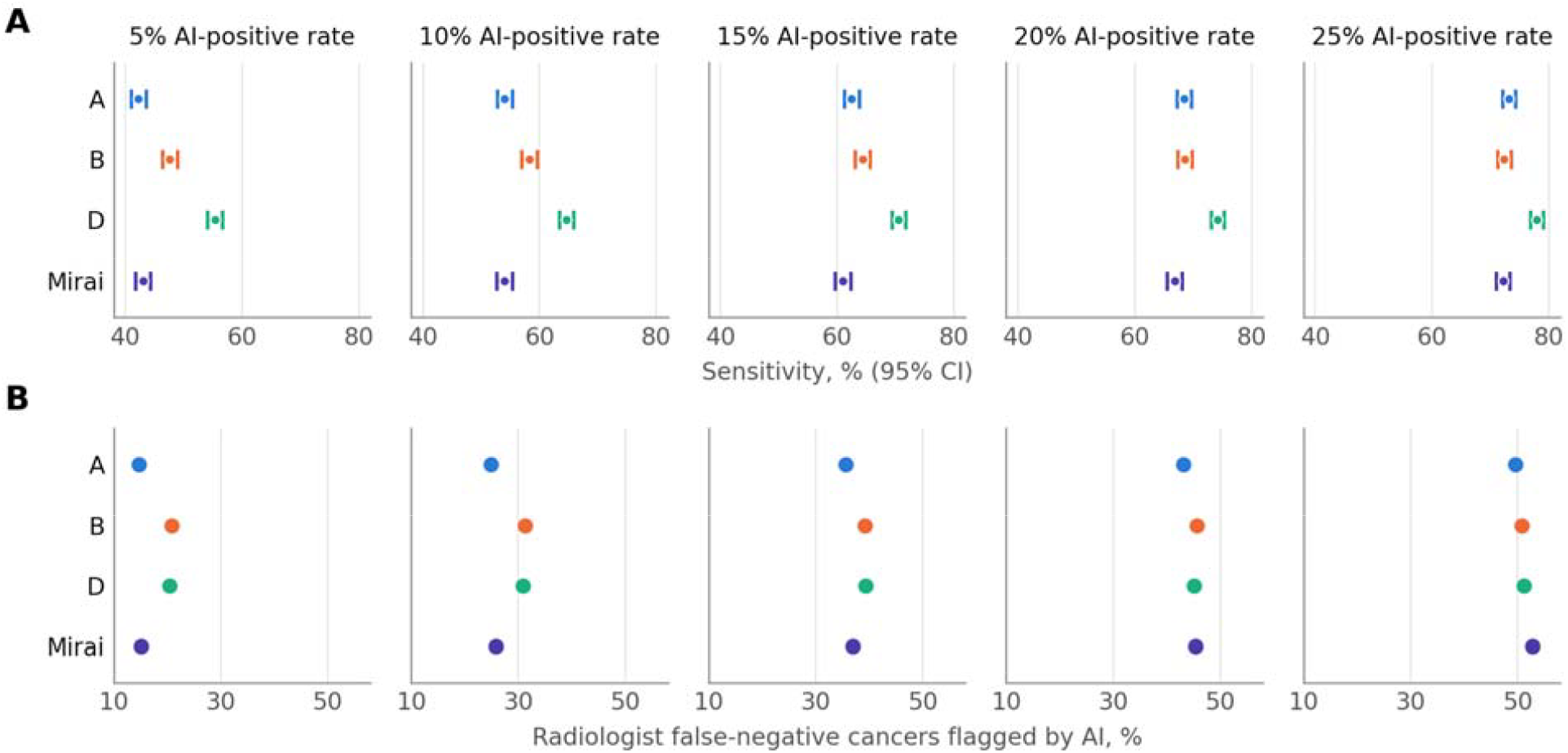
Performance of AI algorithms at matched AI-positive rates of 5%, 10%, 15%, 20%, and 25%. (A) Standalone sensitivity with 95% CIs. (B) Proportion of radiologist false-negative examinations (cancers assessed as BI-RADS 1 or 2 at screening) classified as AI-positive. BI-RADS = Breast Imaging Reporting and Data System.

Algorithm D had the highest sensitivity at every operating point: 55.3% (95% CI: 54.1, 56.6) at 5%, 64.6% (95% CI: 63.4, 65.8) at 10%, 70.5% (95% CI: 69.4, 71.7) at 15%, 74.2% (95% CI: 73.1, 75.3) at 20%, and 77.9% (95% CI: 76.9, 79.0) at 25%, and its 95% CIs did not overlap with those of any other algorithm. Algorithm B ranked second at AI-positive rates of 5%–20% (47.6%, 58.3%, 64.4%, and 68.6%, respectively), with CIs that did not overlap those of algorithm A or Mirai at 5% and 10%. Mirai had sensitivity similar to that of algorithm A at every operating point (43.0% vs 42.2% at 5%; 72.1% vs 73.1% at 25%).

The spread in sensitivity narrowed as the AI-positive rate increased, from 13.1 percentage points at 5% to 5.8 percentage points at 25%. Among algorithms A and B and Mirai, sensitivity ranged from 42.2% to 47.6% at 5% and from 72.1% to 73.1% at 25%, with overlapping CIs at the three highest operating points.

By design, specificity was nearly identical across algorithms at each operating point, ranging from 95.3% to 95.4% at 5% and from 75.3% to 75.4% at 25%. PPV ranged from 6.0% (algorithm A) to 7.9% (algorithm D) at 5% and declined to 2.1%–2.2% at 25%. Achieved AI-positive rates were within 0.1 percentage point of the target for all algorithms.

### Complementarity with Radiologist Interpretation

All algorithms flagged a substantial proportion of the cancers missed by radiologists (Table 3; Figure 2B). At an AI-positive rate of 5%, the algorithms classified 14.7% (algorithm A) to 20.8% (algorithm B) of radiologist false-negative examinations as AI-positive, while flagging 4.1%–4.3% of radiologist true-negative examinations. These proportions increased to 24.9%– 31.3% of false-negative examinations (8.7%–9.0% of true negatives) at 10% and to 49.6%– 52.7% (23.2%–23.6% of true negatives) at 25%. Algorithm A flagged the smallest proportion of radiologist false-negative examinations at every operating point.

**Table 3.** Complementarity of AI Algorithms with Radiologist Interpretation at Matched AI-Positive Rates.

| Algorithm | AI-positive rate |  |  |  |  |
| --- | --- | --- | --- | --- | --- |
|  | 5% | 10% | 15% | 20% | 25% |
| **A. Radiologist false-negative examinations (cancers assessed BI-RADS 1 or 2) classified AI-positive, %** |  |  |  |  |  |
| A | 14.7 | 24.9 | 35.5 | 43.1 | 49.6 |
| B | 20.8 | 31.3 | 39.2 | 45.6 | 50.7 |
| D | 20.5 | 30.8 | 39.3 | 45.1 | 51.1 |
| Mirai | 15.1 | 25.8 | 36.9 | 45.3 | 52.7 |
| **B. Radiologist true-negative examinations (no cancer, BI-RADS 1 or 2) classified AI-positive, %** |  |  |  |  |  |
| A | 4.2 | 8.8 | 13.5 | 18.3 | 23.2 |
| B | 4.3 | 9.0 | 13.8 | 18.6 | 23.6 |
| D | 4.1 | 8.7 | 13.5 | 18.3 | 23.3 |
| Mirai | 4.2 | 9.0 | 13.8 | 18.7 | 23.6 |

Note.—Radiologist assessments were classified as positive (BI-RADS 0) or negative (BI-RADS 1 or 2). BI-RADS = Breast Imaging Reporting and Data System.

The ranking of algorithms for complementarity did not mirror their ranking for standalone sensitivity. Algorithm B flagged a proportion of radiologist false-negative examinations similar to that of algorithm D at every operating point from 5% through 20% (20.8% vs 20.5% at 5%; 45.6% vs 45.1% at 20%), despite its lower standalone sensitivity. At 25%, Mirai, which had the lowest standalone sensitivity at that operating point (72.1%), flagged the largest proportion of radiologist false-negative examinations (52.7%).

In the rule-out analysis (Supplementary Table S1), when the 75% of examinations below the 25% threshold were considered, radiologist-detected cancers were present in 1.6 (algorithm D) to 2.1 (algorithm B and Mirai) per 1,000 AI-negative examinations, and radiologist false-positive recalls accounted for 5.8%–6.3% of AI-negative examinations.

## Discussion

In this national, independent validation of three commercial mammography AI CAD algorithms and the academic Mirai model applied to the same screening examinations in a large and racially and ethnically diverse US population, we found clinically meaningful differences in performance. One commercial algorithm had the highest AUROC and the highest sensitivity at every matched operating point, exceeding the others by as much as 13 percentage points, whereas the other two commercial algorithms performed similarly to Mirai. Every algorithm flagged a substantial fraction of the cancers missed by radiologists, but the algorithms that ranked highest on standalone sensitivity were not always those that best complemented radiologist interpretation.

Our findings extend previous head-to-head evaluations. In a Swedish case-control study, Salim et al found that one of three commercial algorithms significantly outperformed the other two at a matched specificity.^11^ We observed a strikingly similar pattern in a US screening population with single reading, in a cohort in which Asian and Hispanic women together accounted for more than 40% of examinations. The prospective European studies that have established the clinical value of AI-supported screening each evaluated a single product within a double-reading workflow;^5–8^ our results suggest that such findings should not be assumed to transfer to other products or to US practice, where recall rates are higher and the population and equipment differ.

The performance of Mirai is notable. Although it was developed to predict future breast cancer risk rather than to detect prevalent cancer, Mirai discriminated as well as two of the three commercial detection algorithms, consistent with our previous finding that AI algorithms designed for detection and for risk prediction capture overlapping image information.^14^ At the highest operating point, Mirai also flagged the largest proportion of radiologist false-negative examinations, which suggests that image features associated with near-term risk may identify some cancers that are not yet apparent to radiologists. Because Mirai is openly available, it may serve as a reproducible, vendor-independent benchmark against which commercial products can be compared across institutions.

Our results underscore the importance of the operating point. Vendors typically report performance at a recommended threshold, but because sensitivity and specificity trade off against each other, comparisons made at vendor-selected thresholds conflate an algorithm’s discrimination with where its threshold happens to sit. At matched AI-positive rates, specificity and PPV were nearly identical by construction, and sensitivity became the discriminating metric. At an AI-positive rate of 5%, somewhat below the radiologist recall rate of 6.8% in this cohort, no algorithm matched radiologist sensitivity (67.2%), and even at 10%, the sensitivity of the best-performing algorithm (64.6%) remained below that of radiologists. These findings argue against standalone AI interpretation at recall rates comparable to current practice and support the use of AI in combination with radiologists. They also suggest that health systems should establish thresholds locally, based on their own score distributions and their tolerance for additional workup, rather than adopting vendor defaults.

The complementarity analyses are directly relevant to how AI would be deployed. Used as an additional reader, the algorithms flagged 15%–21% of radiologist false-negative examinations while flagging only about 4% of radiologist true-negative examinations at an AI-positive rate of 5%; at 25%, approximately half of radiologist false-negative examinations were flagged. Whether these flags would translate into earlier detection depends on whether the AI finding localizes to the eventual cancer and whether radiologists act on it, questions that require lesion-level and prospective evaluation. Conversely, in a rule-out workflow in which the 75% of examinations with the lowest scores were not read by a radiologist, 1.6–2.1 per 1,000 of those examinations harbored cancers that radiologists had detected, which indicates that autonomous triage cannot be considered safe on the basis of these data. The finding that algorithm rankings differed between standalone sensitivity and complementarity implies that algorithm selection should be tied to the intended clinical use case.

This study had limitations. First, all examinations were two-dimensional digital mammograms; digital breast tomosynthesis now accounts for a large proportion of US screening, and performance on tomosynthesis was not assessed. Second, although the cohort was large and diverse, KP members are insured and may differ from the broader US population; relative to the US female population, Asian women were overrepresented and Black women underrepresented, which limits generalizability to some communities. Third, this was a retrospective evaluation of standalone performance with examinations read without AI; the performance of radiologists working with AI depends on human–AI interaction, which we did not assess. Fourth, AI-positive classification of radiologist false-negative examinations was determined at the examination level and does not confirm that the algorithm localized the cancer. Fifth, the cancer outcome window includes cancers that may not have been visible at the index examination, which would lower the apparent sensitivity of both radiologists and algorithms. Sixth, Mirai was evaluated outside its intended use as a risk model, and we evaluated a single software version of each commercial product, which may be updated over time. Finally, to preserve vendor anonymity, readers cannot link the commercial results to specific products, and subgroup analyses by age, breast density, and race and ethnicity will be reported separately.

In conclusion, commercially available mammography AI algorithms differed meaningfully in performance when evaluated independently on the same US screening examinations at matched operating points, an open-source academic model performed comparably to two of the three commercial products, and each algorithm identified a substantial proportion of the cancers missed by radiologists. Independent, local, multivendor validation at clinically relevant operating points should precede the adoption of mammography AI, and the choice of algorithm and threshold should be matched to the intended clinical workflow.

## Data Availability

All data produced in the present study are available upon reasonable request to the authors

## Supplementary Material

**Supplementary Table S1.** Rule-Out Analysis: Radiologist Outcomes among AI-Negative Examinations.

| Algorithm | AI-positive rate |  |  |  |  |
| --- | --- | --- | --- | --- | --- |
|  | 5% | 10% | 15% | 20% | 25% |
| **A. Radiologist-detected cancers (true positives) among AI-negative examinations, per 1,000** |  |  |  |  |  |
| A | 3.6 | 3.0 | 2.5 | 2.2 | 2.0 |
| B | 3.3 | 2.7 | 2.4 | 2.3 | 2.1 |
| D | 2.7 | 2.2 | 1.9 | 1.8 | 1.6 |
| Mirai | 3.6 | 3.0 | 2.7 | 2.4 | 2.1 |
| **B. Radiologist false-positive recalls among AI-negative examinations, %** |  |  |  |  |  |
| A | 7.1 | 6.7 | 6.4 | 6.1 | 5.8 |
| B | 7.3 | 6.9 | 6.7 | 6.5 | 6.3 |
| D | 7.2 | 6.8 | 6.5 | 6.3 | 6.0 |
| Mirai | 7.2 | 6.9 | 6.7 | 6.5 | 6.3 |

Note.—AI-negative examinations are those scoring below the threshold for the stated AI-positive rate.

